# Modelling the blood bank inventory: simple formulae for stock management in clinical settings

**DOI:** 10.1101/2022.11.04.22281960

**Authors:** S. Vaz, C. Warren, E. Carney, K. Hands, D. Gowans, F. A. Davidson, C. Webb, P. J. Murray

## Abstract

Blood components are a perishable resource that play a crucial role in clinical medicine. The blood component inventory is managed by transfusion services, who ultimately aim to balance supply with demand so as to ensure availability whilst minimising waste. Whilst the blood component inventory problem has been the focus of theoretical approaches for over 50 years, evidence for the direct utilisation of existing models in the day-to-day management of blood stocks in clinical settings is limited. In this study we formulate a discrete mathematical model that describes the main processes in the management of a single population of red blood cells in a clinical setting: ageing, supply and demand. After time averaging the discrete model, a time-delayed integro-partial differential equation model is derived. Steady state analysis yields expressions for: a range of clinically relevant quantities (i.e. age distributions, total stock levels, wastage rates, age of transfused units); key performance indicators; and simple formulae that identify optimal restock thresholds in terms of parameters that are readily available in clinical settings. The approach is validated by testing predictions using data from a Scottish district general hospital. It is envisaged that the proposed methodology can ultimately be used to aid *in situ* ‘rule-of-thumb’ decision making in clinical laboratory settings.

## 1 Introduction

Blood transfusion is an essential part of healthcare worldwide. National transfusion services aim to ensure that there is an adequate supply of red blood cells (RBCs) in individual clinical settings whilst minimising wastage [1, 2]. Packed RBCs represent the major component of transfused blood products. They are routinely transfused for a range of medical treatments (e.g. trauma, surgery, anaemia, cancer treatment) and have a shelf-life of 35 days.

The expression of three surface antigens (A, B and RhD) allows for the categorisation of RBCs into eight major blood groups (see Table 1). For example, Group A^+^ RBCs express A and RhD antigens that would trigger an immune response if transfused into a Group B^+^ patient. In contrast, O^−^ RBCs do not express any of the three surface antigens, and therefore do not usually trigger an immune response when transfused into patients of the other major blood groups. As such, O^−^ RBCs play an essential role in emergency situations when there is insufficient time to determine a patient’s blood group.

**Table 1:** A table illustrating proportions of bloods groups in the UK [3].

| Group | O<sup>+</sup> | O<sup>-</sup> | A<sup>+</sup> | A<sup>-</sup> | B<sup>+</sup> | B<sup>-</sup> | AB<sup>+</sup> | AB<sup>-</sup> |
| --- | --- | --- | --- | --- | --- | --- | --- | --- |
| Percentage of population | 40.9 | 9.5 | 28.8 | 6.3 | 9.2 | 2 | 2.7 | 0.6 |

The supply of packed RBCs is typically regulated by an ‘order-up-to’ restocking strategy. Total stock levels of each blood group are regularly restocked up to threshold levels that are typically established using the experience of transfusion laboratory staff. Additionally, minimal stock thresholds are used to trigger emergency restocks. Restock thresholds tend to be adjusted over time in order to improve performance of the blood inventory.

Demand for RBCs has both predictable and stochastic components. Whilst demand owing to some treatments, such as the support of bone marrow failure or chemotherapy, can be planned for in advance, that owing to, for example, trauma or obstetric haemorrhage is inherently stochastic. To minimise wastage, older units are typically preferentially used, usually via either oldest-unit-first-out (OUFO) or first-in-first-out (FIFO) protocols [2]. Cross grouping of older units can be used to avoid wastage.

At the level of individual clinical settings there have been numerous reports made on features of blood inventories. Key performance indicators (KPI), such as waste as a percentage of issued units (WAPI) [4], issuable stock index (ISI) [5] and the blood-group specific average age of transfused units [6], are used to summarise inventory performance. Moreover, correlations are observed between KPIs: increased restock thresholds are associated with transfusion of older units [7] and higher levels of waste [5]; high levels of cross grouping can also be indicative of issues with inventory performance; and ISI and WAPI are positively correlated [5]).

Significant advances have been made in the modelling and optimisation of the entire blood supply chain (for review see [8]). For example, models of the full blood supply chain have been developed that optimise total cost and age of transfused units [9] and investigate how structural features of supply chains affect overall costs [10]. Numerous studies have also focused on optimisation within individual clinical settings. Simulations of blood bank dynamics have been used for scenario modelling, leading to policy recommendations (e.g. [11, 12]) and demand forecasting has been used to aid in the optimisation of ordering of new stock [13]. Moreover, the effectiveness of different optimisation methods has been studied [14, 15].

A theoretical framework that explicitly relates KPIs to fundamental processes and parameters is thus far limited. In one study, formulae are proposed that relate clinically important quantities (e.g. the minimum age of transfused units, total stock levels) to parameters (e.g. restock frequency) [16]. The potential utility of such an approach in clinical settings is evident in a further study in which the proposed formulae are used to aid the reduction of restock levels so as to reduce the age of transfused units in the inventory [7]. However, as the proposed formulae are derived from heuristic arguments rather than first principles, the extent to which they are a complete description of how fundamental processes (ageing, supply and demand) give rise to observables (e.g. stock levels, waste, average age of transfused units) is unclear.

Despite the significant advances made by previous modelling approaches to the blood inventory problem, evidence of application of theoretical models to the day-to-day management of blood stocks in clinical settings is limited [1]. After interviewing seven transfusion laboratory managers from high-performing transfusion centres in the UK, it was concluded that “in direct opposition to what is claimed in the literature, none of the hospitals surveyed used complex models or equations to readjust target stock levels on a frequent basis” [1].

In this study a simplified model of a blood inventory in a given clinical setting is considered. The population dynamics of a single blood group are considered in a model that characterises supply, demand and ageing. Following derivation of a continuum model, steady state analysis is used to derive closed form expressions that relate key parameters to clinically relevant measurements. The layout is as follows: in Section 2 a model is derived from first principles, in Section 3 model behaviour is explored and a case study using data from a district hospital in the UK; in Section 4 we conclude with a discussion.

## 2 Methods

### 2.1 Model development

#### 2.1.1 A discrete model

Let *t* and *a* be independent variables representing time and age, respectively. Age is discretised with step Δ*a* such that *a* = 0, Δ*a*, …, (*N*_*A*_ − 1)Δ*a* = *A*. Time is discretised with time Δ*t* such that *t* = 0, Δ*t*, …, (*N*_*t*_ − 1)Δ*t* = *T*. Let *Q*_*ij*_ represent the number of units of RBCs in the *i*^*th*^ age interval and *j*^*th*^ time interval.

The ‘order-up-to’ protocol for stock replenishment is captured by a delayed source term with time delay *τ*. Suppose that supply events occur at a set of time indices 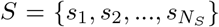. At a given restock event indexed by time *j*,the inventory is restocked to *r* units by adding 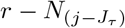 units, where

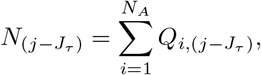

and

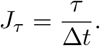

The age distribution of incoming units is determined by sampling from a prescribed discrete probability density function *f*_*i*_, *i* = 1, ‥, *N*_*A*_.

The ‘oldest unit first out’ protocol of stock use is captured by preferentially removing oldest units. Suppose that demand events occur at a set of time indices, 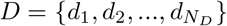. At the *k* demand event, *H*_*k*_ units are removed from the system with oldest units removed until the demand is met.

A model is considered in which ageing, supply and demand are modelled at a set of discrete times and ages. Each supplied unit is represented by an impulse source of random age. Hence

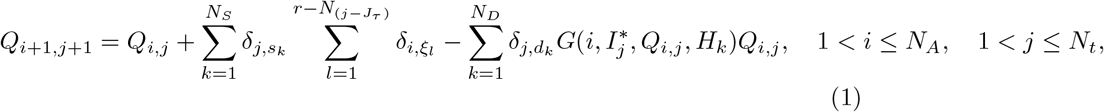

where *δ*_(.,.)_ represents a Kronecker delta function and *ξ*_(.)_ is a randomly sampled age from a prescribed age supply pdf, *f*_*i*_. The OUFO method of stock usage is captured via the function

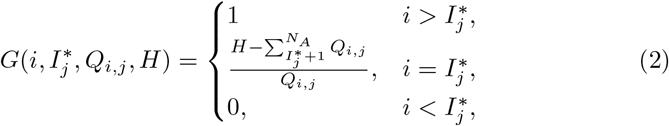

where 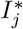, the stock-dependent minimum age of a transfused unit, is computed to be

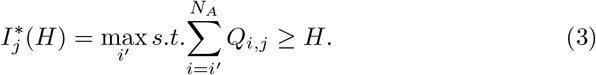

Initial data are given by

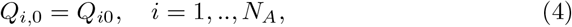

and on the boundary *a* = 0, where there is no flux from a younger age interval,

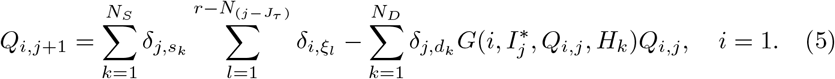

#### 2.1.2 Continuum limit of a time averaged model

We now consider a time-averaged formulation of equations (1)-(5) that yields a limiting continuum model. Consider a time-averaging window of magnitude *T*_*av*_. Discretisation yields

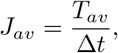

where it is assumed that *J*_*av*_ is a positive integer, and the time-averaged age distribution is given by

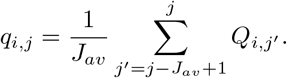

Averaging over equation (1) yields

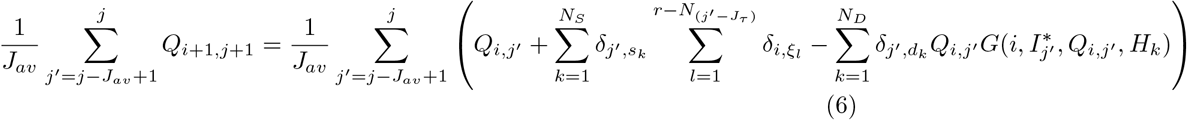

##### Supply term

Suppose that the subset of stocking events that occur in the averaging time interval [*j* − *J*_*av*_ + 1, *j*] is represented by the set of time indices

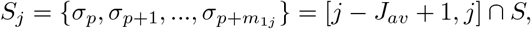

with the number of supply events given by

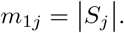

The supply term in equation (6) can be expanded as the sum of *δ*-distributed age distributions

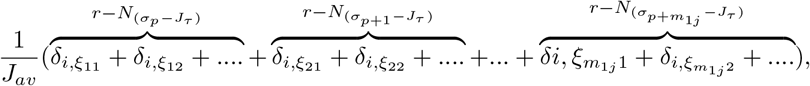

where each of the *ξ*_(.)_ represents a random sample from the prescribed age supply pdf, *f*_*i*_. Gathering terms, the sum can be written as

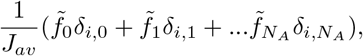

where 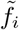 is a random variable representing the total number of supplied units in the *i*^*th*^ age interval. Normalising such that

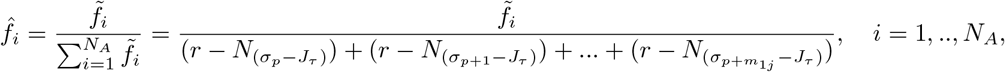

the time-averaged supply term is given by

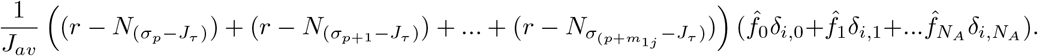

Noting that the sum of Kronecker delta functions is an empirical representation of the age distribution of supplied units, *f*_*i*_, the time-averaged supply term is approximated by

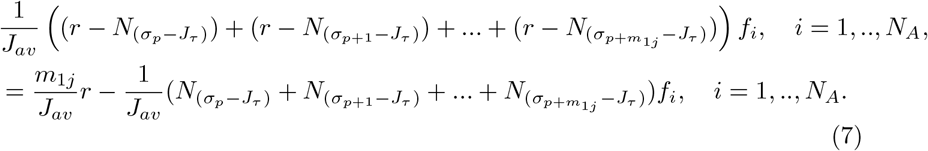

Defining the average total number of units over supply time points to be

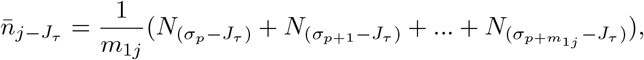

expression (7) can be approximated by the deterministic form

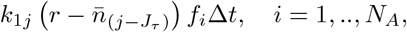

where the time-dependent supply frequency is given by

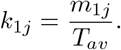

Finally, approximating 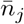 by the average over *J*_*av*_ time points, i.e.

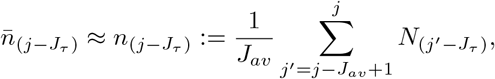

yields source term

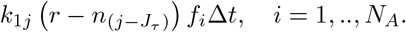

##### Demand term

Suppose that the subset of demand events that occur in the averaging time interval [*j* − *J*_*av*_ + 1, *j*] is represented by the set of time indices

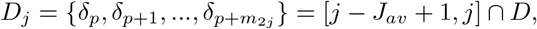

with the number of demand events in the time averaging interval given by

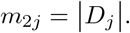

To obtain a tractable limiting model, we approximate that the sum of the individual demand functions can be approximated by

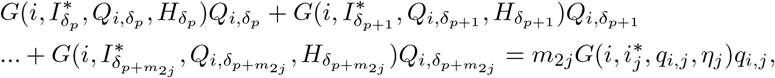

where the time-averaged minimum age of a transfused unit is indexed by

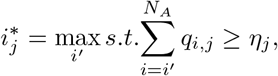

and the time-averaged number of units in demand is given by

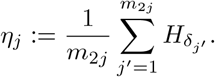

Here, as in the derivation of the supply term, averages computed over the demand instances are approximated by averages computed over all days in the averaging interval. Defining the time-dependent demand frequency

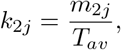

the demand term in equation (6) is approximated by

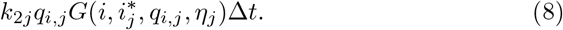

Hence we obtain the discrete time-averaged model

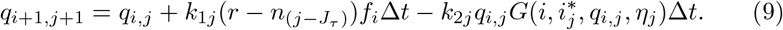

##### The continuum limit

The time-averaged number density is defined to be

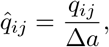

and a continuous age supply pdf, 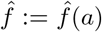, is defined by a point-wise relation to its discrete counterpart:

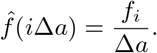

It is assumed that 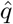 is a continuous function of *a* and *t*. Upon change of variables in equation (9), the limit Δ*t*, Δ*a* → 0 is considered. Upon Taylor expansion, equation (9) can be approximated by the time delayed, integro partial differential equation

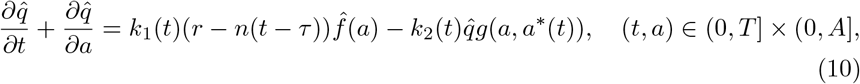

where the total number of units in the inventory, *n*(*t*), is given by

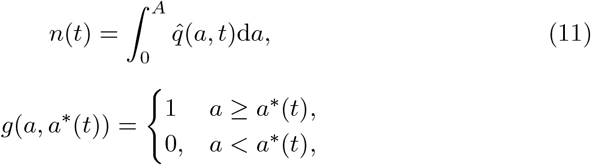

and

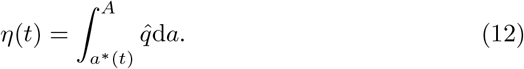

A boundary condition representing no flux at age *a* = 0 is

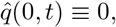

and the initial conditions are

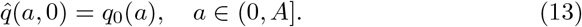

Henceforth, for notational convenience, the hatted notation is omitted.

#### 2.1.3 Steady state

The steady state of equation (10)-(13), 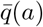, satisfies

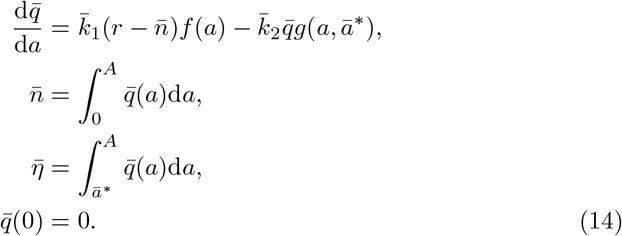

Henceforth, for notational convenience, the barred notation is omitted.

#### 2.1.4 A piecewise linear approximation

To obtain a tractable model, two simplifying assumptions are made regarding the reaction terms in equations (14). It is firstly approximated that the age distribution of supplied units is uniform on the interval [*a*_0_, *a*_1_] where *a*_0_ represents the minimum age of a supplied unit and *a*_1_ is determined by moment matching (see Figure 1). It is further assumed that the expected minimal age of a transfused unit, *a*\*, is greater than *a*_1_, such that supply and demand terms arise in non-intersecting intervals of the age domain ([*a*_0_, *a*_1_] and [*a*\*, *A*], respectively). The validity of the model relies on *a*_0_ *< a*_1_ *< a*\* *< A*, a model assumption that is tested *a posteriori*.

**Figure 1:**
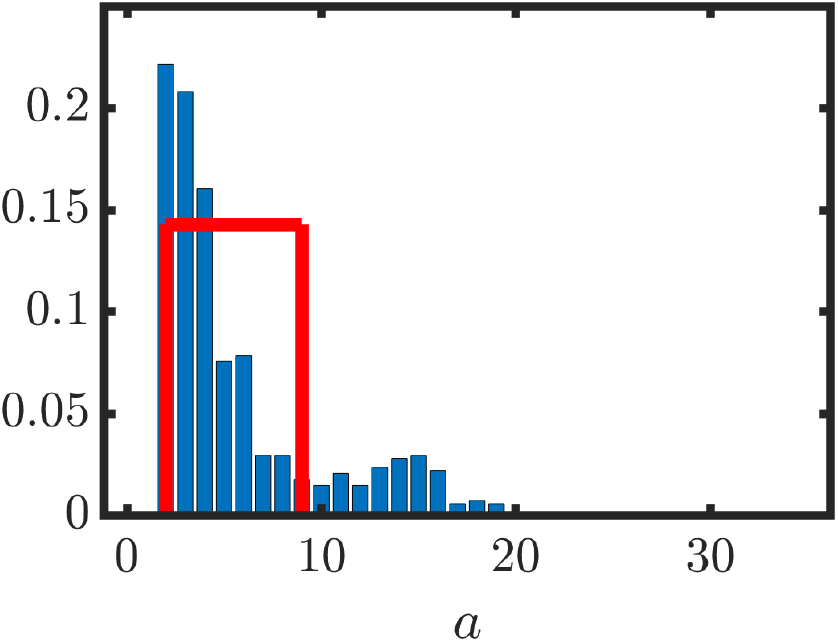
The age (days) distribution, *f*, of supplied units at a Scottish District hospital (blue bars). Red line denotes approximated uniform distribution (see equation (15)). Data provided by Scottish National Blood Transfusion Service.

Suppose new units are supplied to the system at a constant rate *k*_1_ in the age interval *a* ∈ [*a*_0_, *a*_1_]. Hence

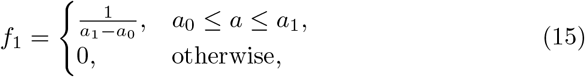

and equations (14) transform to

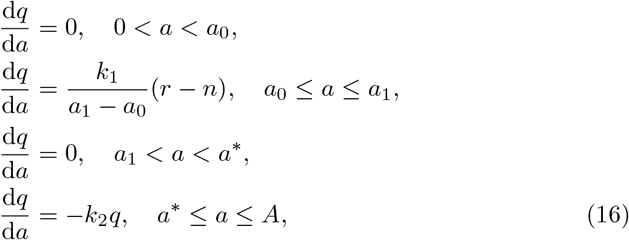

where

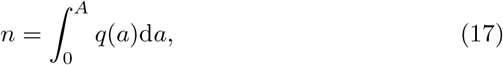

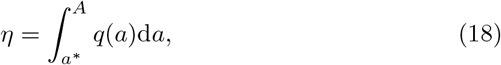

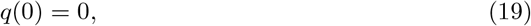

and continuity of *q* is assumed at *a* = *a*_0_, *a* = *a*_1_ and *a* = *a*\*.

After integrating equations (16), application of the boundary and continuity conditions yields a continuous steady state age distribution

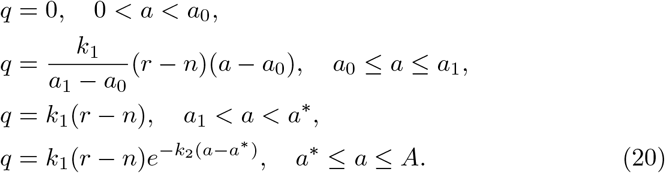

### 2.2 Deriving clinically relevant quantities

#### 2.2.1 Total stock

From equation (17), the total number of units in the inventory, *n*, is given by

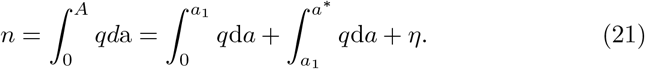

Upon substitution for *q* using equation (20), integration and rearrangement yields

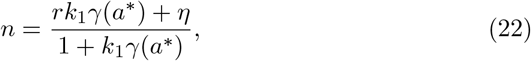

where

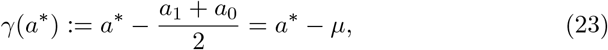

with *µ*, the mean age of a supplied unit:

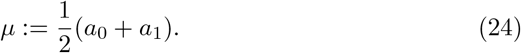

#### 2.2.2 The age of transfused units

##### The minimal age of transfused units

Substituting for *q* using equation (20) in equation (18) yields, after integration and rearrangement, a transcendental equation for the minimum age of issue of a transfused unit

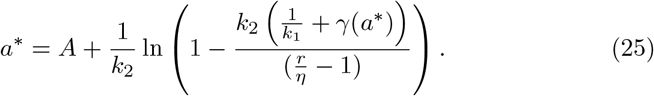

Note that the value of *a*\* is an emergent feature of the model that implicitly depends on model parameters, i.e.

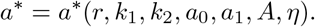

##### An expression for the onset of nonlinearity between *r* and *a*\*

Partial differentiation of equation (25) with respect to *r* yield

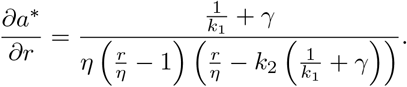

Assuming that *r > η* (trivially satisfied), *a*\* is a strictly increasing function of *r* if

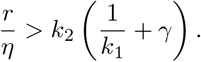

Upon substitution for *r* after rearrangement of equation (25), this inequality is trivially satisfied. Hence for fixed values of parameters *k*_1_, *k*_2_, *η, A* and *µ* there is a one-to-one relationship between *a*\* and r. Upon rearrangement of equation (25)

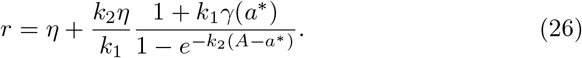

Approximating equation (26) in the limit of small *a*\* (i.e. *a*\* ≪ *A* − 1*/k*_2_) yields

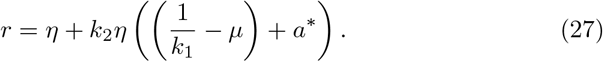

In the limit *a*\* → *A* equation (26) can be approximated by

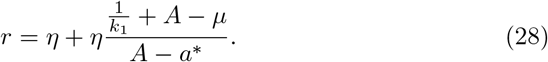

Equating equations (27) and (28) yields an expression for the intersection, *a*_*max*_, (see Figure 2 (d)) that satisfies the quadratic equation

**Figure 2:**
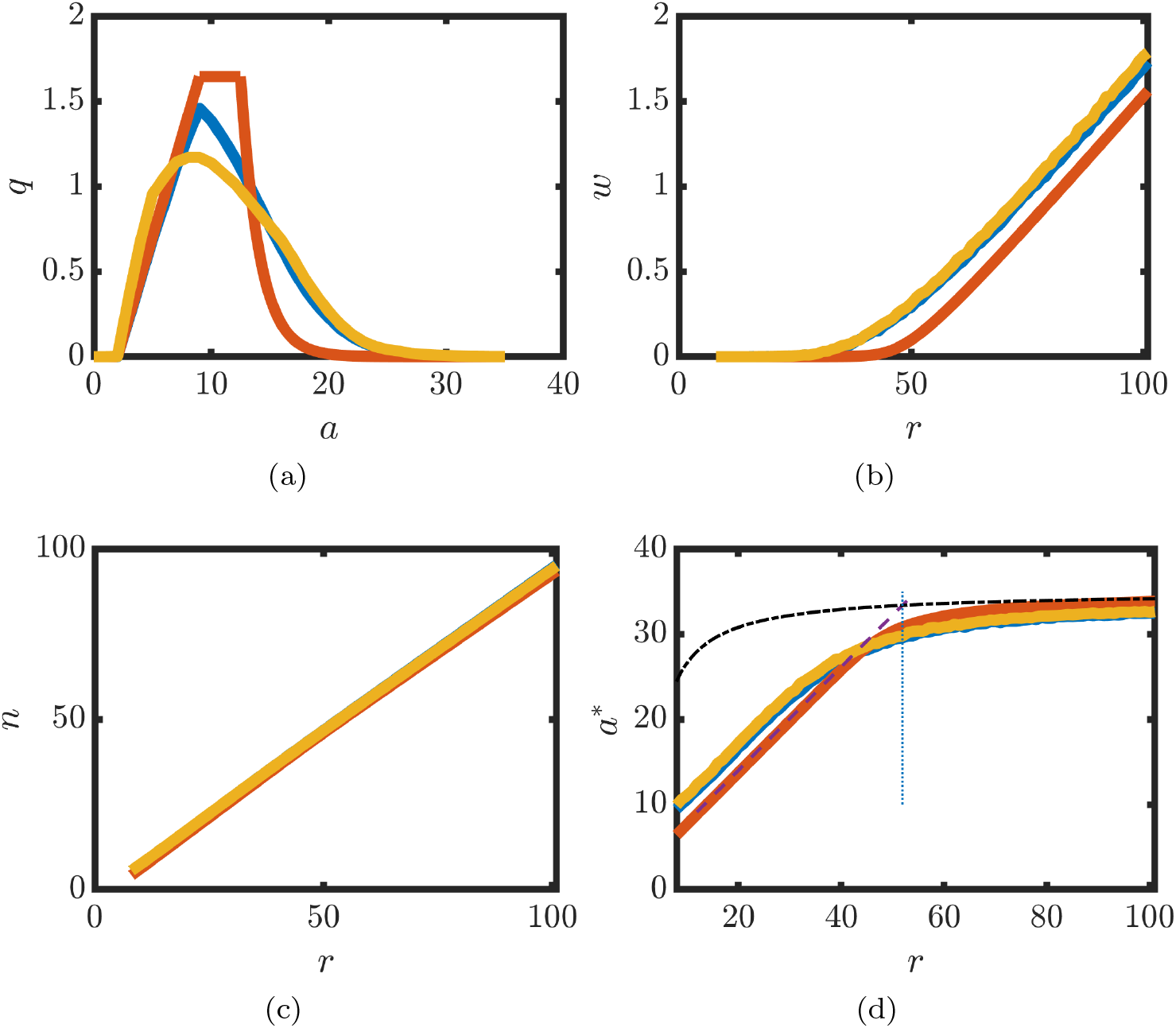
Comparison of key mode outputs. (a) Number density, *q*, is plotted against age, *a*. *r* = 18. (b) The wastage rate, *w*, is plotted against restock threshold, *r*. (c) Total stock, *n*, is plotted against restock threshold, *r*. (d) Minimum age of transfused units, *a*\*, is plotted against restock threshold, *r*. Blue lines - time average of discrete model with uniformly distributed age supply pdf. Yellow lines - time average of discrete model with empirically estimated age supply pdf. Red lines - continuum model. Continuum model solutions for *q, w, n* and *a*\* given by equations (20), (33), (22) and (25), respectively. Asymptotes in (d) given by equations (27) and (28). Dotted vertical lines represent onset of waste (equation (30)). *k*_2_ = 1.57. *τ* = 0. Discrete model given by equations (1)-(5). Other parameters as in Table 3.

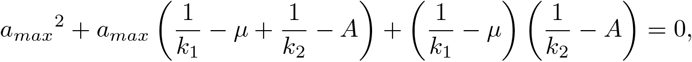

and has root

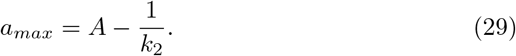

Substitution in equation (27) yields

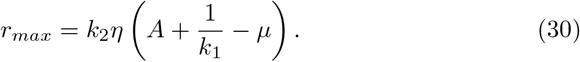

Note that the other root

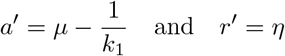

yields an inadmissible solution (*a*\* *< µ < a*_1_) and is therefore omitted in the analysis below.

##### An expression for the expected age of transfused units

The expected age of transfused units, a quantity that can be readily compared with available data, is given by

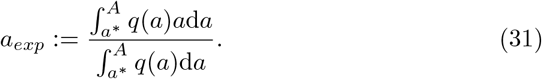

After substitution for *q* using equation (20) and integration,

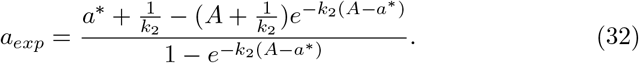

#### 2.2.3 Waste

The wastage rate, *w*, represents the loss of units as a result of reaching the expiry age, *A*. Upon substitution for *a* = *A* in the last equation in (20), it follows that

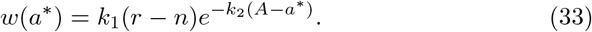

Rearranging equations (22) yields

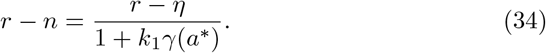

Substitution for equation (26) in the right-hand side of equation (34) yields

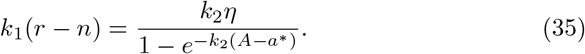

Finally, upon substitution in equation (33), we obtain

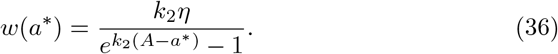

#### 2.2.4 WAPI

The waste as a percentage of issued units (WAPI) is a KPI that is used to quantify the level of waste in an inventory [5]. In the proposed model the net supply rate is

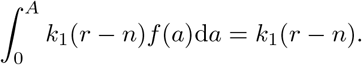

Hence the WAPI is represented by

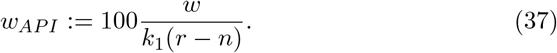

Upon substitution for *w* using equation (33) and cancellation

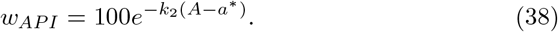

#### 2.2.5 ISI

The issueable stock index (ISI) is a KPI that quantifies the number of days of issueable stock in an inventory. It is computed to be the ratio of the number of unreserved red cell units to the net supply rate [5]. In the model the ISI is therefore represented by

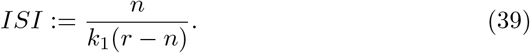

Substituting for *n, r − n* and *γ* using equations (22), (34) and (23), respectively, yields

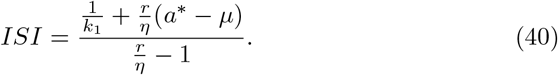

Notably, the steady state wastage rate, WAPI, ISI and expected age of transfused units can be expressed as functions of model parameters (with implicit dependence via *a*\* using equation (25)).

#### 2.2.6 The low wastage limit

Equation (36) implies that when *k*_2_(*A* − *a*\*) ∼ 1, the wastage and transfusion rates are approximately equal (i.e. approximately 50% of units are wasted). As WAPI scores in practice are of the order a few percent [5], transfusion laboratories operate in the low wastage limit:

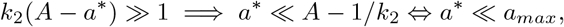

where *a*_*max*_ defined by equation (29). In this limit, equation (32) can be approximated by

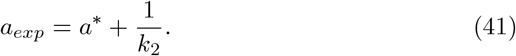

Moreover, inversion of the relationship between *a*\* and *r* given by equation (27) yields

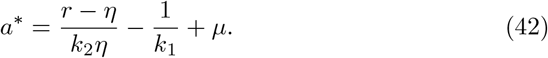

Thus an explicit expression for the minimum age of transfused units is identified in the low wastage limit.

Upon substitution for *a*\* using equation (42) in equations (38), (40) and (41), expressions for the WAPI, ISI and expected age of transfused units, respectively, in the low wastage limit are given by

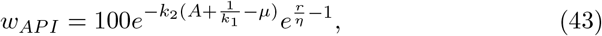

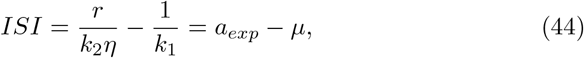

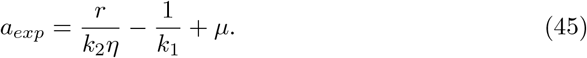

Moreover, the condition for model validity *a*\* *> a*_1_ can be expressed, upon substitution using equation (27), as

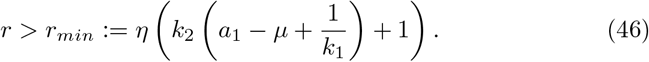

### 2.3 Optimisation

#### 2.3.1 Minimising waste and maximising stock

To identify optimal restock thresholds, an objective function, *E*, is defined that penalises wastage and low stock. Consider

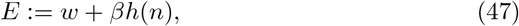

where *w* is waste as defined above and *h*(*n*) is a decreasing function of total stock levels, with *β* a constant that describes relative importance of low stock. As *β* characterises the trade off between waste and stock depletion in a given clinical setting, it is expected that it will vary depending on factors such as management policy, geographical location etc. Here we consider a linear penalty for low stock levels is represented by

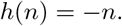

Substitution for *w* and *n* in equation (47) yields, after some calculation,

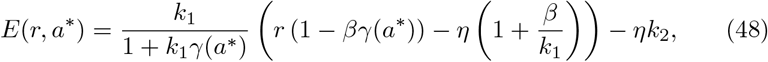

with *a*\* define implicitly via equation (25). Ultimately, we wish to identify the value of the restock threshold, *r*, that minimises E given the implicitly defined *a*\* via equation (25).

Using equation (27) to substitute for *r* in equation (48) yields

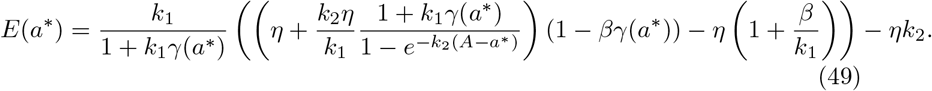

Hence one can seek a value of *a*\* that minimises equation (49) and obtain the corresponding *r* using equation (26). The minimiser of equation (49) is denoted by *r*_*E*_.

#### 2.3.2 Explicit formulae for target KPIs in the low wastage limit

Suppose that a transfusion laboratory wishes to obtain a target WAPI of *T*_*W API*_, a target ISI, *T*_*ISI*_ or a target expected age, *T*_*age*_. Substitution for the target values in equations (43), (44) and (45), respectively, yields, upon rearrangment, corresponding restock thresholds *r*_*W API*_, *r*_*ISI*_ and *r*_*age*_ given by

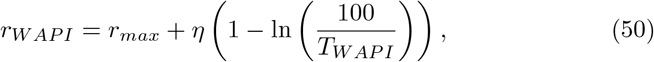

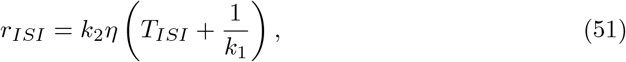

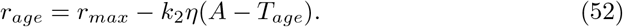

### 2.4 Numerical solutions

#### 2.4.1 Simulation

To simulate equation (1) we considered Δ*t* = Δ*a* = 1. Stocking times were chosen at days *S* = 1, 3, 5, 8, 10, 12, … to represent restocking on Mondays, Wednesdays and Fridays. The waiting time between demand events was sampled from a geometric distribution with mean waiting time 1*/k*_2_, i.e. the probability of a unit demand event happening on a given day is

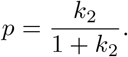

Hence the set of days, *D*, on which demand events was identified. On each demand day, *η*_*k*_ units were removed. An initial condition

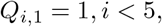

was considered.

On restocking days the total number of units was computed in order to determine the number of units that are needed. Two possibilities were considered for the age of supplied units: (i) uniform sampling from the age interval [*a*_0_, *a*_1_]; or (ii) sampling from an empirical estimate of the pdf obtained from Figure 1. On a demand day, the oldest units in the system are removed until demand is satisfied. A burn-in time of 50 days was used to allow the system to reach dynamic equilibrium before summary statistics are computed. The wastage rate was computed to be the number of units on the boundary *i* = *N*_*A*_ at the end of each time step.

#### 2.4.2 Steady state age distribution

To compute the steady state age distribution (equation (20)) *a*\* is firstly computed by numerically computing the root of the transcendental equation (25). This was achieved in Matlab using the root-finding algorithm ‘fsolve’.

### 3 Results

### 3.1 Model exploration

To explore the dynamics of blood stocks in clinical settings, a mathematical model (equations (1)–(5)) was developed that describes the supply, demand and ageing of units of a single blood group. The supply term is modelled using the ‘order-up-to’ principle by which the total stock measured at some delay time *t* − *τ* determines the number of units supplied at time *t*, with the age of each supplied unit sampled from the uniform distribution [*a*_0_, *a*_1_]. The demand term, which captures an oldest unit first out (OUFO) protocol for stock management, is also non-local as oldest units are removed until demand is satisfied. The model is stochastic as the age of supplied units and time of demand events are random variables.

The discrete model (equations (1)–(5), see Section 2 2.4) was simulated for long times and time-averaged, steady-state age distributions were computed (see Figure 2 (a)). Key clinically measurable quantities (i.e. total stock, *n*, wastage rate, *w*, and the minimum age of transfused units at different) were sampled at different values of restock threshold, *r* (see Figure 2 (b)-(d)). The results indicate that, for the chosen parameters, total stock levels depend linearly on restock threshold, *r*, whilst both the wastage rate, *w*, and the minimum age of transfused units, *a*\*, exhibit biphasic dependence on *r*. The minimal age of a transfused unit, *a*\*, is an increasing function of restock threshold *r* (see Figure 2 (d)): a consequence of stocking more units is that patients tend to receive older units. When *r* is small (see Figure (2) (b) and (d)), the system behaves like a ‘just in time’ supply chain: transfused units have only recently been supplied and waste is negligible. In contrast, when *r* is large, *a*\* is close to maximal age *A*; units are held in the inventory until just prior to their expiry and the wastage rate is large. Notably, the time averaged solutions did not vary significantly when an empirical estimate of the age supply pdf was used (i.e. with either of the supply age distributions depicted in Figure 1; see the blue and yellow curves in Figure 2 (a)).

Whilst direct simulation approaches can provide summary information on how a model behaves for a specific set of parameters (e.g. Figure 2), an inherent limitation is that they cannot readily provide qualitative descriptions of how model behaviours depend on model parameters (e.g. how does wastage rate depend on *r* given a different restocking frequency *k*_1_?). To address this issue the discrete model (equations (1)–(5)) was time averaged (see Section 2 2.1.2), yielding a continuum limit that is a time-delayed, age-structured, integro partial differential equation model (see Section 2 2.1.2). The continuum model (equations (10)-(13)) has a relatively small number of parameters, all of which can be readily determined in a given clinical setting. The parameter *k*_1_ represents the average frequency of supply events whilst the age distribution of supplied units is captured by the parameters *a*_0_ and *a*_1_. The parameter *k*_2_ represents the frequency of demand events (i.e. blood transfusions) and will vary in proportion with the prevalence of particular blood groups in the population (see Table 2). The parameter *r* represents the ‘order-up-to’ restock threshold of a particular blood group.

**Table 2:** A table summarising blood-group dependent parameters, predicted KPIs and estimated restock thresholds. *AB*^+^, *AB*^−^ and *B*^+^ were not stocked. Top: Time period 1. Bottom: Time period 2. See Section 2 2.2 for derived quantities.

|  | O<sup>+</sup> | O<sup>-</sup> | A<sup>+</sup> | A<sup>-</sup> | B<sup>-</sup> |
| --- | --- | --- | --- | --- | --- |
| $r_{Used}$ | 18 | 12 | 14 | 6 | 2 |
| $k_2$ | 1.57 | 0.50 | 1.07 | 0.24 | 0.15 |
| $W_{API}$ | 0.00 | 0.79 | 0.00 | 8.18 | 2.25 |
| $ISI$ | 10 | 22 | 10 | 23 | 11 |
| $r_{min}$ | 10 | 4 | 7 | 2 | 2 |
| $r_{max}$ | 50 | 16 | 34 | 8 | 5 |
| $r_E$ | 44 | 11 | 28 | 3 | 1 |
| $r_{WAPI}$ | 45 | 11 | 29 | 2 | 0 |
| $r_{age}$ | 20 | 6 | 14 | 3 | 2 |
| $r_{ISI}$ | 19 | 6 | 13 | 3 | 2 |
| $r_{Used}$ | 18 | 8 | 14 | 4 | 2 |
| $k_2$ | 1.62 | 0.51 | 1.18 | 0.38 | 0.26 |
| $W_{API}$ | 0.00 | 0.00 | 0.00 | 0.001 | 0.07 |
| $ISI$ | 9 | 13 | 10 | 8 | 5 |
| $r_{min}$ | 11 | 4 | 8 | 3 | 3 |
| $r_{max}$ | 52 | 16 | 37 | 12 | 8 |
| $r_E$ | 45 | 11 | 31 | 7 | 4 |
| $r_{WAPI}$ | 46 | 11 | 32 | 7 | 3 |
| $r_{age}$ | 21 | 7 | 15 | 5 | 3 |
| $r_{ISI}$ | 20 | 6 | 15 | 5 | 3 |

The steady-state age distribution (equation (20)) is linear in the supply age interval (i.e. *a*_0_ *< a < a*_1_) and exponentially decaying in the demand age interval (see Figure 2 (a)). Using the steady state age distribution, expressions can be derived for: the wastage rate (equation (33)), total stock level (equation (22)), and the expected age of transfused units (equation (41)). Excellent agreement was observed between steady states of continuum and corresponding discrete model quantities (see Figure 2 and Appendix A). Steady state solutions of the model also readily yield expressions for KPIs [5]: the waste as a percentage of issued units (WAPI), a KPI that computes the wastage rate as a percentage of the supply rate (see Section 2 2.2.4), is given by

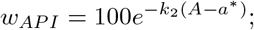

and the ISI (see Section 2 (2.2.5)), a KPI that estimates the number of days of stock in an inventory, is given by

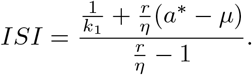

These expressions, in which *a*\* depends implicitly on the model parameters via the transcendental equation (25), indicate that WAPI and ISI are exponentially and linearly increasing functions of the minimum age of transfused units, respectively. This result is consistent with observed positive correlation between WAPI and ISI measured across multiple clinical settings [5].

Explicit expressions can be derived for KPIs in the low wastage limit. Notably, the response of *a*\* to *r* is approximately biphasic (see Figure 2 (d)); a value of *r* at which transition occurs (see equation (30)) is estimated to be

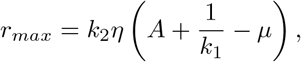

with the corresponding value of *a*\* given by

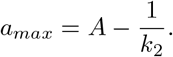

Furthermore, *r*_*max*_ represents the stock threshold at which the wastage rate is approximately equal to the transfusion rate (i.e. as many units are being wasted as are being used, see Section 2.2.3). As WAPI scores are of the order a few percent in clinical settings [5], it is expected that transfusion laboratories operate in the low wastage limit (i.e. *r* ≪ *r*_*max*_ ⟺ *a*\* ≪ *a*_*max*_). In the model this implies that the minimum age of transfused units can be approximated by (see equation (27))

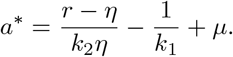

Hence, explicit forms for the WAPI, ISI and expected age of transfused units are given by (see Section 2 2.2.6)

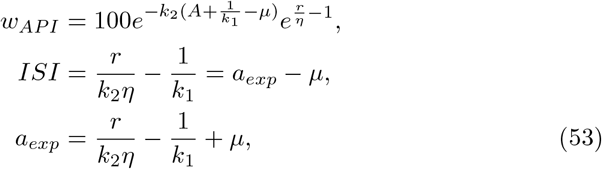

respectively. These derived forms provide insight into how key performance indicators relate to model parameters. For example, the WAPI increases exponentially with restock thresholds whilst the ISI increases linearly. Although care must taken when comparing observations taken from different clinical centres, as multiple parameters are likely different, the derived formulae are consistent with observed positive correlation between WAPI and ISI metrics [5].

Upon inversion of equations (53), the restock threshold that yields a target WAPI, *T*_*W AP I*_, target ISI, *T*_*ISI*_ or target mean age of transfused units, *T*_*age*_, are given by

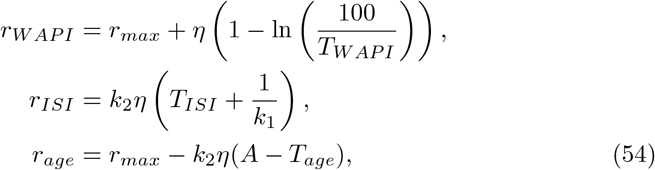

respectively.

### 3.2 Application of the model in a clinical setting

To validate the proposed model, blood transfusion records from a Scottish district general hospital (DGH) were obtained. Over the one year period for which data were provided by the Scottish National Blood Transfusion Service (SNBTS), restocking occurred on Monday, Wednesday and Friday. Hence the average restocking frequency, *k*_1_, was determined. Using the known age distribution of supplied units (see Figure 1), the parameter *a*_0_ was identified as the minimum age of a supplied unit and *a*_1_ was determined using equation (24) and matching with mean of the empirical distribution. Using the total number of transfused units of each blood group, the blood-group specific demand rates, *k*_2_, were identified (see Table 2). Notably, over the time period during which the data were obtained, the restock thresholds, *r*, were modified (thresholds for *O*^−^ and *A*^−^ were reduced), yielding a natural experiment in which the predicted effects of perturbing thresholds can be validated (see Table 2). In each of the time periods, the mean age of transfused units at the DGH was computed and compared with the model prediction in the low wastage limit (see Figure 3 and equation (41)). The results suggest that the model can accurately predict how the expected age of transfused units varies across different blood groups. Moreover, the model also captures dependence of the expected age of transfused units on restock thresholds (e.g. note decrease mean age of transfused unit as a result of decreasing *O*^−^ restock thresholds).

**Figure 3:**
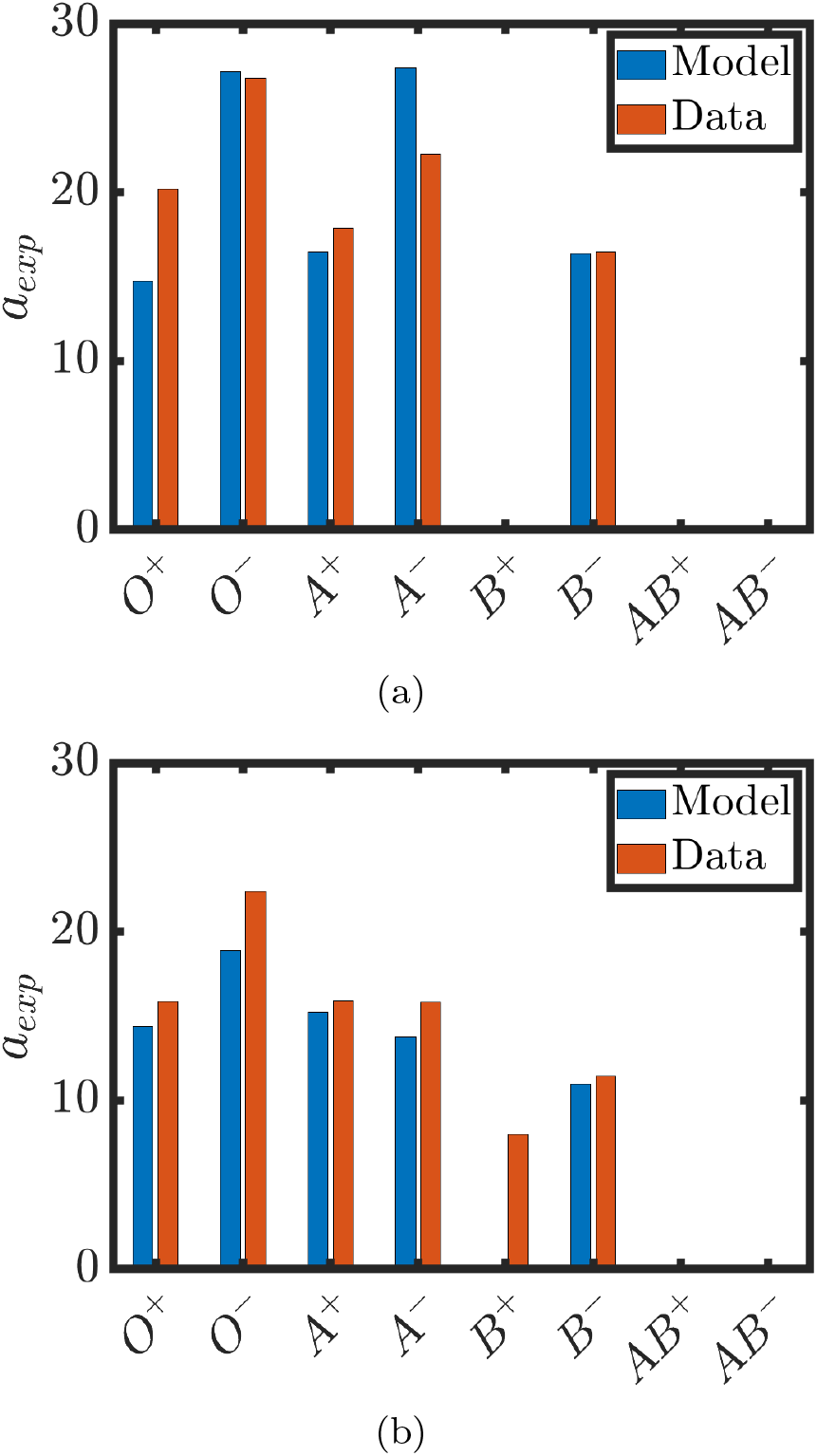
Expected age of transfused units, *a*_*exp*_, is plotted for different blood groups. (equation (41)). (a) Restock threshold for time period 1. (b) Restock thresholds for time period 2. Model data (blue bars) generated using equation (41). See Tables 2 and Table 3 for parameter values.

The model was used to predict values of the key performance indicators: WAPI and ISI. The predicted WAPI was less than 0.7% for each blood group (see Table 3). Notably, values of WAPI of the order of 3% have been reported in the literature [5]. However, in the time period of study, there was zero wastage reported at the DGH under study. The model was also used to compute predicted ISIs (national averages are reported to be approximately 6 days [5]). It is notable that in Time Period 1 two blood groups (*O*^−^ and *B*^−^) had predicted ISIs that were markedly larger than the others (see Table 2). Subsequently, the thresholds for these blood groups were reduced and the predicted ISIs in Time Period 2 then aligned with those of the other blood groups. These results suggest that, for the dataset under study, the predicted ISI could be used to aid rule-of-thumb decision making.

**Table 3:** A table of model parameters.

| Parameter | Definition | Value | Unit |
| --- | --- | --- | --- |
| $T$ | Simulation end time | 2000 | d |
| $\Delta t$ | Simulation time step | 1 | d |
| $A$ | RBC expiry age | 35 | d |
| $k_1$ | Supply frequency | 0.43 | d<sup>-1</sup> |
| $\mu$ | Mean age of supplied units | 5.65 | d |
| $a_0$ | Min. age of supplied units | 2.0 | d |
| $\beta$ | Optimisation weight | 0.001 | d<sup>-1</sup> |
| $T_{WAPI}$ | Target WAPI | 0.2 | nondim |
| $T_{ISI}$ | Target ISI | 10 | d |
| $T_{age}$ | Target expected transfusion age | 16 | d |

To explore how model solutions depend on restock thresholds for the baseline parameter sets at the DGH under study, the minimum age of transfused units, *a*\*, (see Figure 4 (a)) and waste (see Figure 4 (b)) were computed over a range of values of restock threshold, *r*, for each blood group. As expected, the restock threshold at which the wastage rate is approximately equal to the transfusion rate (i.e. *r*_*max*_) is significantly larger than the used restock thresholds (compare values of *r*_*max*_ and *r*_*Used*_ in Table 2); the transfusion laboratory operates in the low wastage limit.

**Figure 4:**
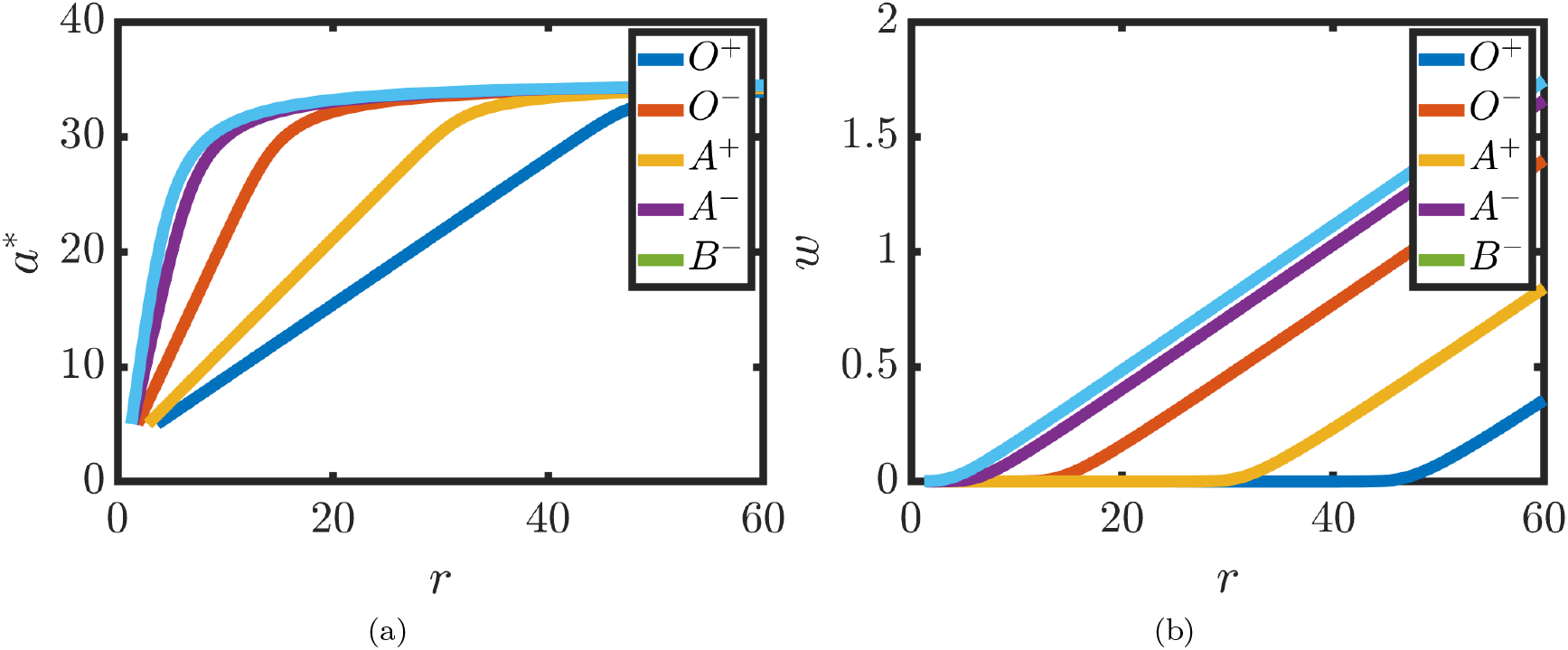
Exploring the effect of varying the restock threshold, *r*. (a) The minimum age of transfused units, *a*\*, is plotted against *r* (see equation (41)). (b) Wastage rate, *w*, is plotted against restock threshold, *r*, (equation (33)). Parameter values (for Time period 1) as in Tables 2 and 3 unless otherwise stated. Note that *B*^+^, *AB*^+^ and *AB*^−^ were not stocked.

To explore whether the used thresholds are ‘optimal’, an objective function, *E*, was constructed that accounts for the balance of two of the major constraints faced in clinical settings (to minimise wastage whilst guarding against (critically) low levels of stock - see equation (47)). Values of minimal transfusion age, *a*\*, were identified that minimise the objective function for each of the blood groups over a range of values of the weighting parameter *β* (see Section 2 2.3.1 and Figure 5 (a)-(c) for representative examples). It was found that the optimal value of restock threshold, *r*_*E*_, was reasonably stable over a range of values of *β* (Figure 5 (d)-(f)). Interestingly, it was found that the restock thresholds that minimise the objective function are significantly larger than those used for blood groups A^+^ and O^+^ (see Table 2). The results suggest that these blood groups could be stocked at higher levels without incurring wastage.

**Figure 5:**
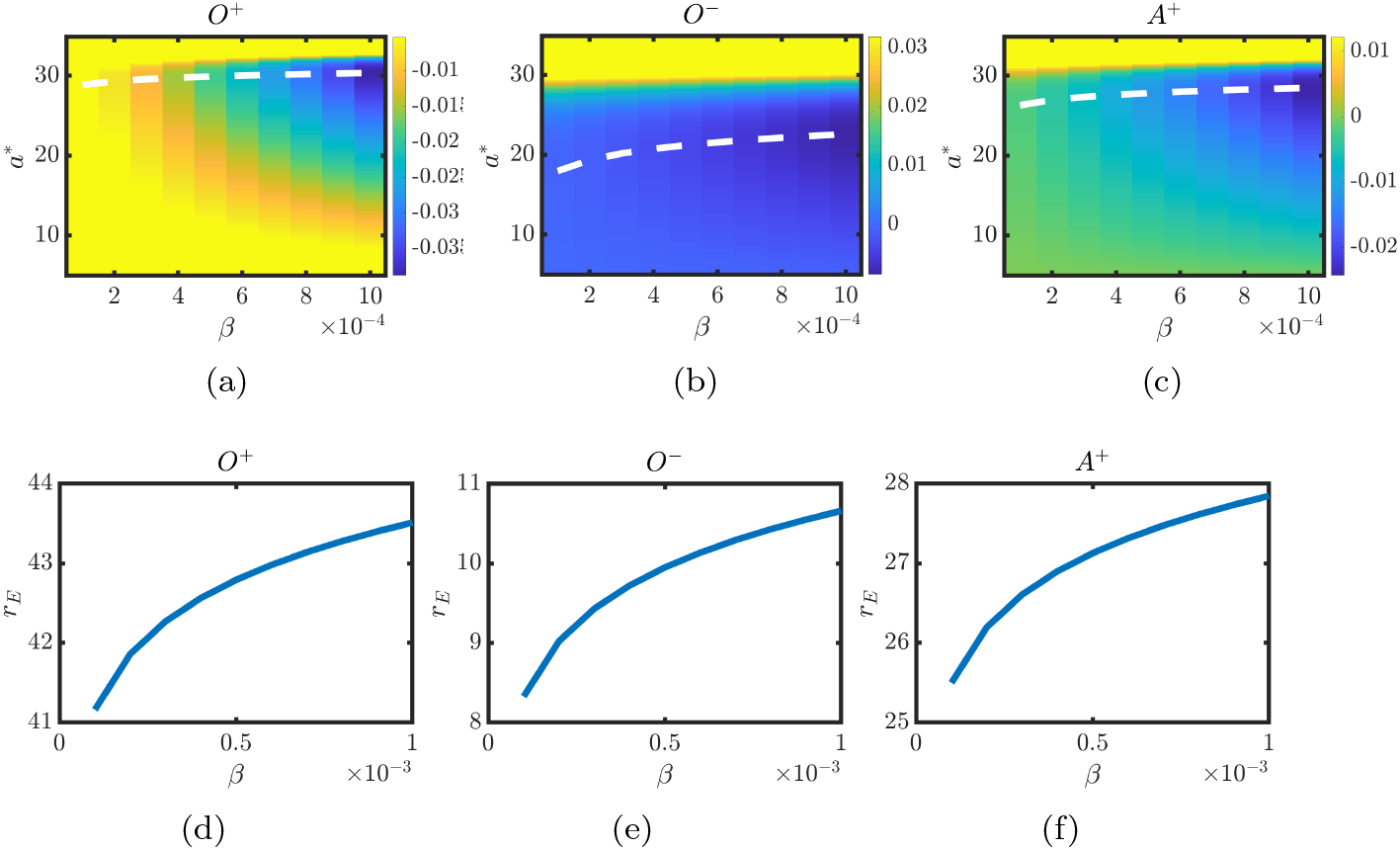
Optimal restock thresholds for selected blood groups. (a-c) The value of the objective function, *E*, (see equation (48)) is plotted against, *a*\*, and *β*. Colour represents *E* and the dashed lines local minima. (d-f) Optimal solution, *r*_*E*_, is plotted against weighting parameter, *β* (*r*_*E*_ is computed by numerically finding *a*\* that minimises *E* and then using equation (26)). Parameter values as in Tables 2 and 3 unless otherwise stated.

Alternatively, upon using the target formulae presented in equation (54), it was found that: setting *T*_*W API*_ = 0.2 provides similar estimates of restock thresholds to those obtained using numerical optimisation of equation (47) (see Figure 5); choosing *T*_*age*_ ∼ 16 provides a good estimate for the threshold values used at the DGH over the time period of interest (see Figure 6); and setting *T*_*ISI*_ = 10 yields a reasonable estimate for used restock thresholds. These results suggest that the proposed model could be used to aid rule-of-thumb decision making for the dataset under study.

**Figure 6:**
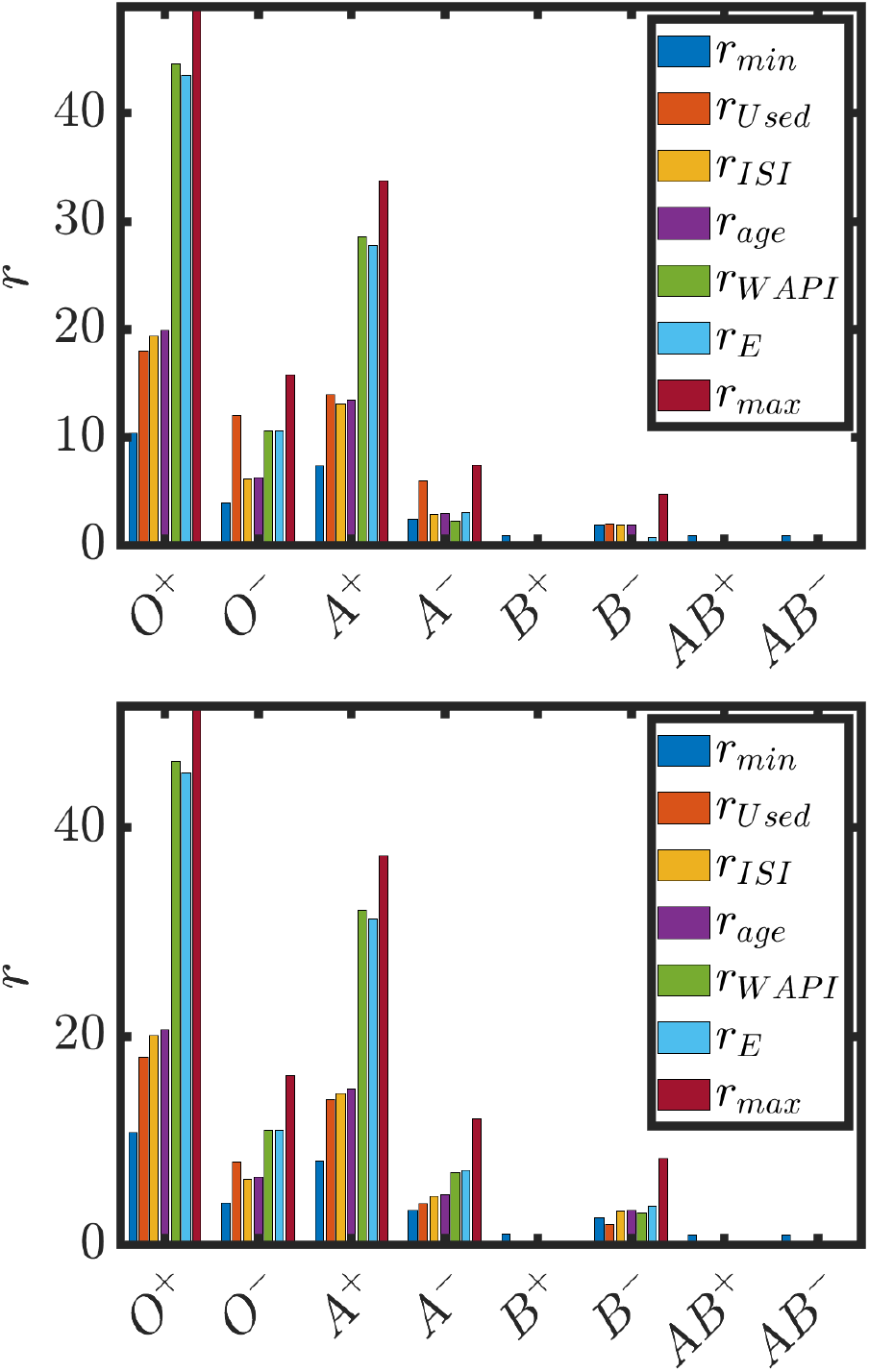
A summary of different estimates for restock thresholds for the different blood groups. (a) Time period 1. (b) Time period 2. Parameter values as in Tables 2 and 3. *T*_*W AP I*_ = 0.2, *T*_*age*_ = 16.

## 4 Conclusion

As blood transfusions are required for many clinical treatments (e.g. surgery, childbirth, chemotherapy), the availability of blood products plays a vital enabling role in modern clinical medicine. Blood inventories are managed by transfusion laboratories who, ultimately, manage two opposing constraints: minimise waste whilst simultaneously ensuring that adequate stock is available to meet clinical demand. Whilst numerous theoretical approaches have been applied to the blood inventory problem, their direct impact on day-to-day management of blood stocks in transfusion laboratories is limited. There is therefore a need for complementary modelling approaches that yield outputs that can aid day-to-day decision making in transfusion laboratories.

In this study a discrete stochastic model of individual blood groups was considered that accounts for ageing, supply and demand of RBC units in an inventory. Upon time averaging and taking a continuum limit, an age-structured, time-delayed, integro-partial differential equation was obtained. An explicit form was found for the steady state solution which was then used to derive expressions for a number of clinically relevant quantities. Analysis of the steady state solutions indicates that both the wastage rate and the mean age of transfused units have a biphasic dependence on the restock threshold, *r*, and an expression was derived for the restock threshold, *r*_*max*_, at which the wastage rate is approximately equal to the demand rate. It was assumed that transfusion laboratories choose restock thresholds to be significantly less than this upper bound (i.e. *r* ≪ *r*_*max*_) and hence that inventories operates in the limit of low wastage. The steady state solution of the model allows for predictions of KPIs in terms of known model parameters. The forms for predicted KPIs are consistent with observations of correlations between KPIs. Moreover the derived forms for the KPIs can be inverted, yielding formulae for restock thresholds that will yield target values of KPIs. Hence the model yields simple formulae that could aid rule-of-thumb decision making in clinical laboratories.

A case study was considered in which the model was applied to data from a Scottish DGH. Data were obtained from two distinct time periods within which different restock thresholds were used. In each of the time periods the model captures variation in the mean age of transfused across the different blood groups [6]. Moreover, the model captures the change in mean age of transfused units upon reduction of two of the restock thresholds.

An optimisation problem was constructed in which an objective function that penalises both waste and low stock levels was minimised. The identified ‘optimal solutions’ suggest that *O*^+^ and *A*^+^ could be stocked at increased levels than was the case at the DGH under consideration. In the model, the effect of the increased restock thresholds was to increase stock levels with a minimal effect on wastage. It is noted that supply side constraints on stock availability are a potential reason for using lower restock thresholds (even if wastage may not be increased at higher stock values).

Model-predicted ISIs were used to retrospectively diagnose and address an issue with the inventory. When explicit formulae that yield target of values of ISIs were used, the model retrospectively identified predicted ISIs for two blood groups that were significantly higher than the other blood groups. The stock thresholds for the identified two blood groups were then reduced by the transfusion laboratory and the predicted ISIs, computed using the modified restock thresholds, were aligned with those of the other other blood groups.

The management of blood inventories will vary dependent on a host of site-specific factors (e.g. geographical proximity to blood bank, demand rates, trauma unit). For example, in a DGH located in close proximity to a central blood bank with frequent routine deliveries and few trauma cases, it may be desirable to have lower restock thresholds than a DGH in a remote geographical location. It is noted that whilst the derived formulae for KPIs provide a means of systematically comparing the behaviour of multiple blood inventories, further study, that considers transfusion records from multiple clinical settings, is required to determine the general applicability of the proposed model in its current form.

Whilst the model provides insight into features of the blood inventory problem, a number of modelling assumptions could be revisited in order to refine the model predictions. Firstly, when taking the continuum limit it is assumed that supply and demand are distributed homogeneously in time. In reality, demand frequency is structured in time (i.e. there are strong circadian and weekly time scales). Moreover, one also expects day-to-day age structure in the supply of units as, ultimately, units are provided by donors and this tends to happen on particular days of the week (e.g. there will be relatively few donations on a Sunday).

It is noted that the continuum model exhibits a steeper gradient in the steady state age distribution than the time-averaged discrete simulations (see Figure 2 (a)) and that the continuum model underestimates both the mean age of transfused units and the wastage rate (see Figure 2 and Appendix A). The source of the deviation is an assumption made in the derivation of the demand term in the continuum model in which the time average of Heaviside functions is itself a Heaviside function (see equation (8)). These factors ought to be taken into account when comparing derived formulae with clinical records.

In general, the demand for a given blood group is coupled to levels of other blood groups in the system (i.e. cross grouping). Cross-grouping is particularly important in the context of the management of *O*^−^ stocks, as O^−^ units are routinely transfused to patients of any blood group in emergency situations. Moreover, cross-grouping can be used as a management strategy to avoid wastage. Whilst the model in its current form does not directly describe cross-grouping, the effect of cross grouping is manifest via the demand rates for each blood group (which are calculated by averaging the overall demand (includes homologous and cross grouped transfused units)).

The modelling approach adopted in this study provides formulae that allow the value of restock thresholds to be identified that yield target values of KPIs. A major advantage of our approach is that the derived formulae can be readily computed given available knowledge of statistics of the particular supply and demand rates in a given clinical setting. Whilst the derived formulae are in a form that could be readily used to inform rule-of-thumb decisions made by transfusion laboratory staff, further data are needed to establish the general validity across different clinical settings.

## Data Availability

All data reported in the present work are contained in the manuscript.

## Appendix A

To further investigate agreement between discrete and continuum models the wastage rate and mean age of transfused units were computed over a range of values of restock threshold, *r*, and demand frequency, *k*_2_ (see Figure 7). Note that the discrete and continuum models show excellent quantitative agreement over a wide range of values of restock threshold and demand frequency. However, the continuum model underestimates the wastage rate in the case where *a*\* ∼ 30 and it underestimates the minimum age of transfused units, *a*\* as 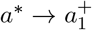.

**Figure 7:**
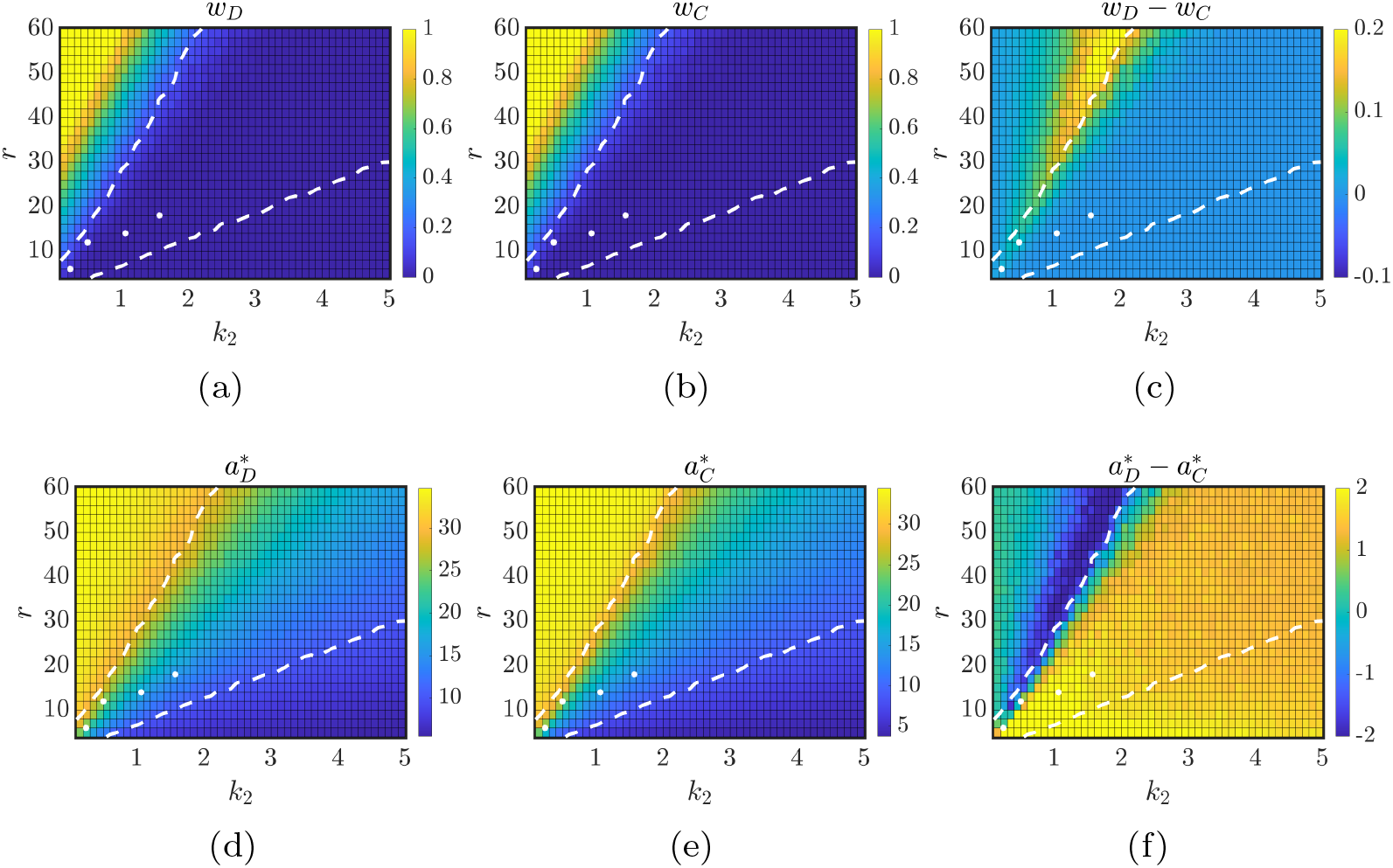
Comparison of discrete and continuum model outputs. Wastage rate, *w*_*D*_, in the discrete model is plotted against restock threshold, *r*, and demand rate, *k*_2_. (b) The corresponding wastage rate in the continuum model, *w*_*C*_. (c) The difference between wastage rates in the discrete and continuum models (*w*_*D*_ −*W*_*C*_). (d) The minimum age of transfused units in the discrete model, 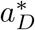, is plotted against restock threshold, *r*, and demand rate, *k*_2_. The corresponding minimum age of transfused units in the continuum model, 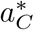. (f) The difference between minimum age of transfused units in the discrete and continuum models 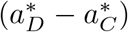. Dashed lines represent contours *a*\* = *a*_1_ and *a*\* = 30. White dots represent parameters used for different blood groups at the Scottish DGH under study (see Table 2). Continuum model solutions for *w* and *a*\* given by equations (33) and (25), respectively. *τ* = 0. Discrete model given by equations (1)-(5). Other parameters as in Table 3.

## Notes

### Competing Interest Statement

The authors have declared no competing interest.

### Funding Statement

The authors acknowledge funding via a DCAT vacation scholarship to CW and an Edinburgh Mathematical Society student research bursary to EC.

### Summary of Updates

updated document style.

## References

[1] Stanger SH, Yates N, Wilding R, Cotton S. 2012 Blood inventory management: hospital best practice. Transfusion Medicine Reviews 26, 153–163.

[2] Williamson LM, Devine DV. 2013 Challenges in the management of the blood supply. The Lancet 381, 1866–1875.

[3] Scot Blood. https://www.scotblood.co.uk/about-blood/blood-types/. Accessed: 2020-09-21.

[4] Yates N, Stanger S, Wilding R, Cotton S. 2017 Approaches to assessing and minimizing blood wastage in the hospital and blood supply chain. ISBT Science Series 12, 91–98.

[5] Perera G, Hyam C, Taylor C, Chapman J. 2009 Hospital blood inventory practice: the factors affecting stock level and wastage. Transfusion Medicine 19, 99–104.

[6] Owens W, Tokessy M, Rock G. 2001 Age of blood in inventory at a large tertiary care hospital. Vox sanguinis 81, 21–23.

[7] Poisson JL, Tuma CW, Shulman IA. 2016 Inventory management strategies that reduce the age of red blood cell components at the time of transfusion. Transfusion 56, 1758–1762.

[8] Osorio AF, Brailsford SC, Smith HK. 2015 A structured review of quantitative models in the blood supply chain: a taxonomic framework for decisionmaking. International Journal of Production Research 53, 7191–7212.

[9] Zahiri B, Torabi SA, Mohammadi M, Aghabegloo M. 2018 A multi-stage stochastic programming approach for blood supply chain planning. Computers & Industrial Engineering 122, 1–14.

[10] Osorio AF, Brailsford SC, Smith HK, Blake J. 2018 Designing the blood supply chain: how much, how and where?. Vox sanguinis 113, 760–769.

[11] Katsaliaki K, Brailsford SC. 2007 Using simulation to improve the blood supply chain. Journal of the Operational Research Society 58, 219–227.

[12] Mohammadi N, Seyedi SH, Farhadi P, Shahmohamadi J, Ganjeh ZA, Salehi Z. 2022 Development of a scenario-based blood bank model to maximize reducing the blood wastage. Transfusion Clinique et Biologique 29, 16–19.

[13] Fortsch SM, Khapalova EA. 2016 Reducing uncertainty in demand for blood. Operations Research for Health Care 9, 16–28.

[14] Adewumi A, Budlender N, Olusanya M. 2012 Optimizing the assignment of blood in a blood banking system: some initial results. In 2012 IEEE congress on evolutionary computation pp. 1–6. IEEE.

[15] Duan Q, Liao TW. 2014 Optimization of blood supply chain with shortened shelf lives and ABO compatibility. International Journal of Production Economics 153, 113–129.

[16] Dzik WH, Beckman N, Murphy MF, Delaney M, Flanagan P, Fung M, Germain M, Haspel RL, Lozano M, Sacher R et al. 2013 Factors affecting red blood cell storage age at the time of transfusion. Transfusion 53, 3110– 3119.

